# Understanding of vascular toxicity of indigo naturalis in anti-inflammation application: mendelian randomization and colocalization study

**DOI:** 10.1101/2025.08.07.25332888

**Authors:** Jie Liu, Cheng Jian Liu, Hai Tao Xiao, Zhi Ping Xu

## Abstract

The Chinese herbal medicine indigo naturalis (IN) shows promising potential for treating autoimmune inflammatory diseases. However, its clinical application is limited by vascular toxicity. Using Mendelian randomization analysis, we conducted the first systematic study of IN’s target-specific vascular risks, revealing causal links between IN exposure and various vascular disorders. Importantly, our findings identify urate as a mediator between IN exposure and deep vein thrombosis outcome. This study provides genetic insights into safety implications for clinical translation study of IN for anti-inflammation applications and guides biomarker monitoring in related trials.

## 1. Introduction

Indigo naturalis (IN, Qing Dai in Chinese) is a well-established anti-inflammation herb of traditional Chinese medicine (TCM) for the treatment of autoimmune inflammatory disorders including psoriasis and ulcerative colitis (UC).^1^ As such, its clinical translation has been attracting considerable research effort,^2–6^ yet being hurdled by the poor understanding on its vascular toxicity. In multiple clinical trials and practices, the toxicity typically manifested in the form of portal vein thrombosis (PVT)^4^ and pulmonary arterial hypertension (PAH),^7, 8^ but the underlying mechanism remains elusive. In this context, a comprehensive understanding of the serious toxicity is in demand to facilitate the translation study of IN.

The bis-indole alkaloid indigo (ING) and its isomer indirubin (INB) are the major organic components of IN, at least 4% and 0.13%, respectively, and recognized as the active pharmaceutical ingredients (APIs) against psoriasis and ulcerative colitis by TCM expert consensus.^1^ ING and ING are both potent agonists of aryl hydrocarbon receptor (AhR) participating in immunological and metabolic regulations.^1^ Moreover, INB is the antagonist of three types of kinases, i.e., fms like tyrosine kinase 3 (FLT3),^9,10^ cyclin-dependent kinases (CDKs) and glycogen synthase kinase-3β (GSK-3β).^1, 1–13^ So, we propose that the vascular toxicity of IN is associated with the targets of ING and INB and the reasoning is given as follows. First, both alkaloids can elicit pro-inflammatory effect by activating AhR/CYP1A1 metabolic axis^8, 14^ and subsequently promoting reactive oxygen species (ROS) production and NLRP3 activation.^15^ Second, INB can particularly cause inflammation due to its potent cytotoxicity through inhibiting CDKs and GSK-3β.^1, 11–13,16^ In fact, INB has been clinically used to kill leukemia cells since 2002 for the treatment of chronic myelocytic leukemia in China.^17^ Moreover, INB can disrupt certain vascular repair functions through deactivating FLT3 given that FLT3 mediates differentiation of hematopoietic progenitor cells^9^ and development and maintenance of dendritic cells.^10^ Taken together, these risk factors suggest that exposure of INB/ING of IN would cause the vascular toxicity through interacting with their targets, i.e., AhR, CDKs, GSK-3β and FLT3. To date, the evidence supporting the hypothesis remains limited to one preclinical study on the toxic effect of AhR activation by IN.^8^

As a prominent methodological alternative to randomized controlled trials (RCTs), Mendelian randomization (MR) has been emerging as a promising tool for drug safety prediction in pharmaceutical development,^18, 19^ with its key advantages in substantially reducing biases from reverse causation and residual confounding by using genome-wide association study (GWAS) data.^20, 21^ Briefly, MR enables causal inference between drug exposure and toxicity risks by leveraging genetic variants associated with both gene expression of drug targets (e.g., AhR, CDKs, GSK-3β, and FLT3) and phenotypic outcomes (e.g., PVT and PAH).

To the end, this work will use MR to investigate that whether changes in gene-expression levels of ING and INB’s targets (i.e., AhR, CDK1-20, GSK-3β and FLT3) may causally affect vascular risk. In this study, it is hypothesized that there may be a causal effect of expression levels of AhR, CDK1-20, GSK-3β and FLT3 on 30 of vascular risks. Using 2-sample MR and genetic colocalization analyses, we found that expressions of AhR, FLT3, CDK4, 6 and 16 support a causal effect (*P*<0.05) on multiple vascular risks. Furthermore, mediation MR indicated that urate mediates one vascular risk (i.e., deep venous thrombosis, DVT) of IN exposure by AhR activation. This work for the first time comprehensively provides MR evidence of causal effects of APIs of IN (i.e., ING and INB) on vascular toxicity.

## 2. Methods

### 2.1 Study design and dataset description

As primary analyses MR was used to examine the following: the effects of gene-expression levels of ING/INB targets on vascular risk. Upon identifying MR evidence of a causal effect, colocalization analysis was conducted to confirm that the exposure and outcome were regulated by the same causal variant. Mediation effect was further investigated by screening metabolites, immune cells and gut bacteria.

All GWAS summary datasets were obtained from IEU GWAS database as listed in Table S1, including 30 of vascular conditions and 23 of quantitative trait loci (eQTL) of gene expressions (AhR, FLT3, GSK-3β, CDK1-20).

### 2.2 Selecting drug targets

Gene targets for ING and INB were collected from references.^12, 13^ The eQTL datasets of the target genes (AhR, FLT3, GSK-3β, CDK1-20) is listed in Table S1.

### 2.3 Two-sample MR and mediation MR analyses

The inverse variance weighted (IVW) MR method was selected as our main MR analysis method. Two-sample MR was performed by MR-Base platform (*P* = 5×10^−5^),^22^ where Mendelian Randomization Pleiotropy Residual Sum and Outlier (MR-PRESSO) was used. Mediation effect was examined by screening metabolites, immune cells and gut bacteria (*P* = 5×10^−5^) with GWAS summary datasets in IEU GWAS database.^23^

### 2.4 Sensitivity analyses

Several sensitivity analyses were performed to determine the presence of pleiotropy and heterogeneity. Pleiotropy analysis was mainly based on the MR-Egger intercept test and the heterogeneity test using Cochran’s Q statistic. The analyses showed statistically significant differences when P < 0.05.

### 2.5 Colocalization analysis

Colocalization analysis was performed using R package Coloc (version3.2-1).^24^ Variants ± 100kb of reference variant were included. The 1000 Genomes v3 European ancestry dataset was used as LD reference panel. Evidence for colocalization was defined as a posterior probability greater than 0.8.

## 3. Results

### 3.1 Effect of ING/INB targets on vascular risk

MR was conducted with 30 of vascular conditions as outcomes and 23 of ING/INB targets (AhR, FLT3, GSK-3β, and CDK1-20) as exposure at the *P* = 5×10^−5^ level (Figure 1a). MR evidence of causal effect of expressions of AhR, FLT3, and CDK4/6/14 on 5 of vascular disorders was found, respectively (Table 1 and Table S1-6).

**Table 1.**
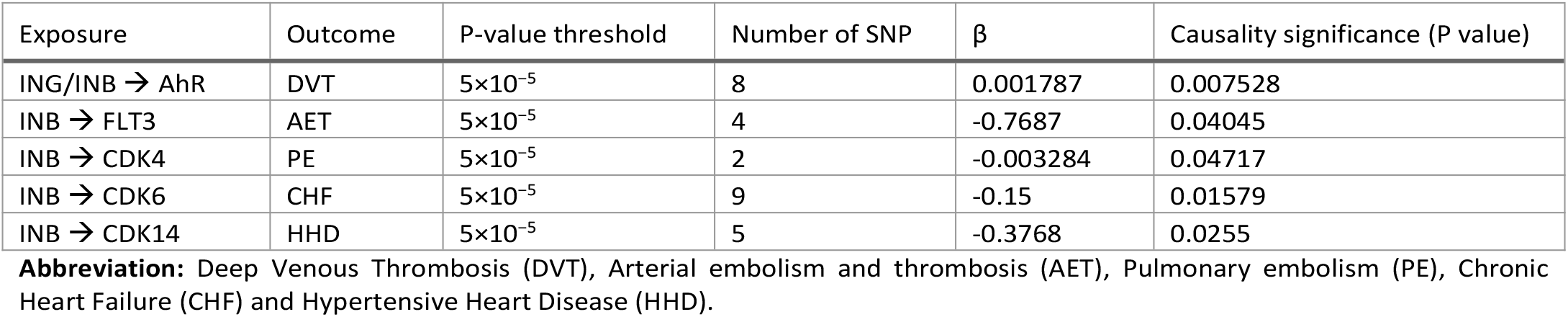
Results of the MR analyses with ING/INB as exposures and vascular risks as outcomes (*P* = 5×10^−5^)

| Exposure | Outcome | P-value threshold | Number of SNP | $\beta$ | Causality significance (P value) |
| --- | --- | --- | --- | --- | --- |
| ING/INB → AhR | DVT | $5 \times 10^{-5}$ | 8 | 0.001787 | 0.007528 |
| INB → FLT3 | AET | $5 \times 10^{-5}$ | 4 | -0.7687 | 0.04045 |
| INB → CDK4 | PE | $5 \times 10^{-5}$ | 2 | -0.003284 | 0.04717 |
| INB → CDK6 | CHF | $5 \times 10^{-5}$ | 9 | -0.15 | 0.01579 |
| INB → CDK14 | HHD | $5 \times 10^{-5}$ | 5 | -0.3768 | 0.0255 |

**Abbreviation:** Deep Venous Thrombosis (DVT), Arterial embolism and thrombosis (AET), Pulmonary embolism (PE), Chronic Heart Failure (CHF) and Hypertensive Heart Disease (HHD).

**Figure 1.**
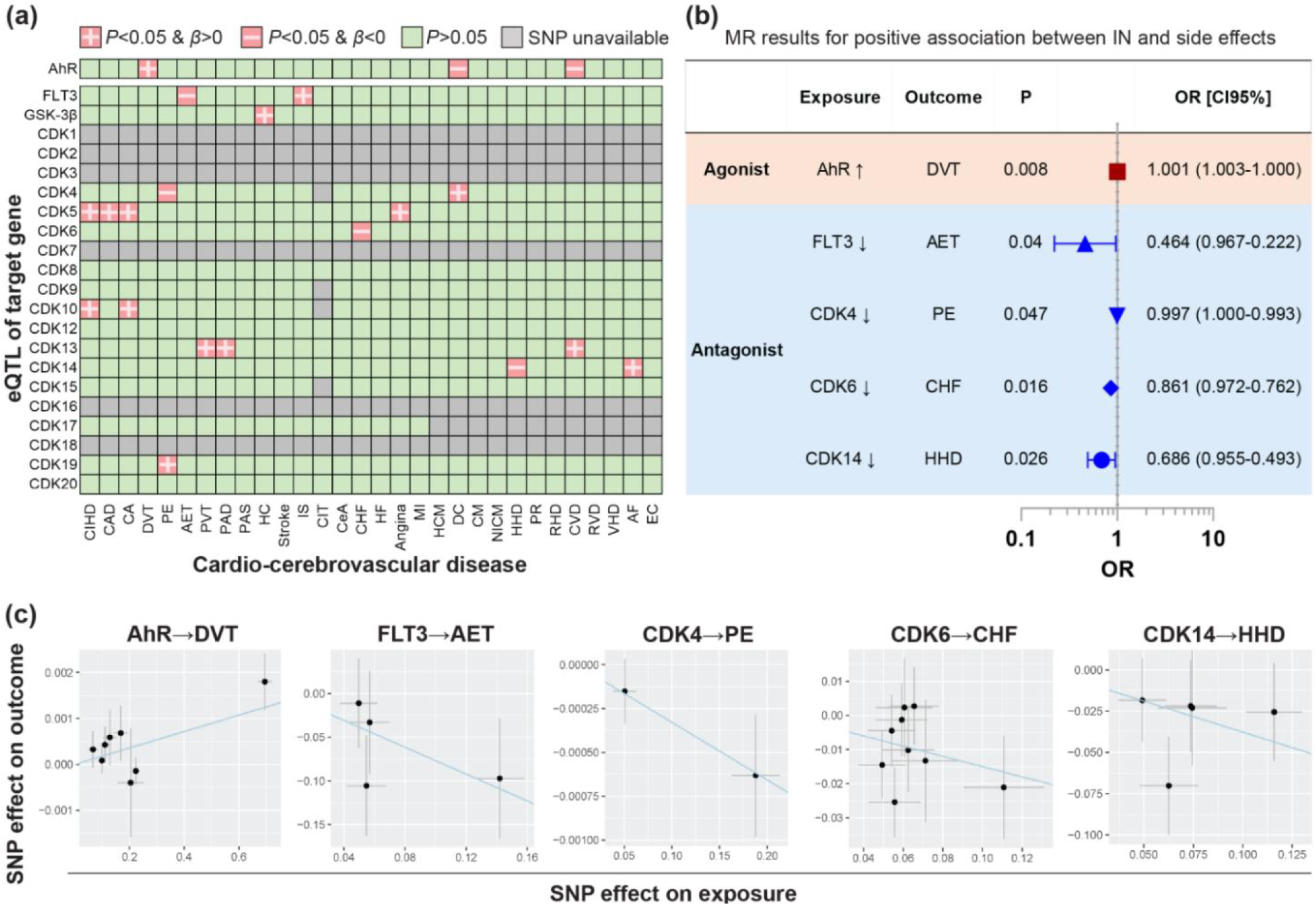
MR results at the *P* = 5×10^−5^ level. (a) Heatmap of MR results with eQTLs of ING/INB target genes (exposure) and 30 of vascular diseases (outcome); (b) Summary of MR results indicative of a causal effect of ING/INB exposure on vascular toxicities; (c) Scatter plot of MR results. **Abbreviation:** Chronic ischaemic heart disease (CIHD), Coronary artery disease (CAD), Coronary atherosclerosis (CoA), Deep venous thrombosis (DVT), Pulmonary embolism (PE), Arterial embolism and thrombosis (AET), Haemmorrhoids and perianal venous thrombosis (PVT), Peripheral artery disease (PAD), Peripheral atherosclerosis (PAS), High cholesterol (HC), Stroke (Stroke), Ischemic stroke (IS), Carotid Intima-media thickness (CIT), Cerebral atherosclerosis (CeA), Chronic heart failure (CHF), Heart failure (HF), Angina (Angina), Myocardial infarction (MI), Hypertrophic cardiomyopathy (HCM), Dilated cardiomyopathy (DC), Cardiomyopathy (CM), Nonischemic cardiomyopathy (NICM), Hypertensive Heart Disease (HHD), Polymyalgia rheumatica (PR), Rheumatic heart disease (RHD), Cardiac valvular disease (CVD), Rheumatic valve diseases (RVD), Valvular heart disease (VHD), Atrial fibrillation (AF), and Endocarditis (EC).

Specifically, MR demonstrates a positive effect of AhR expression on deep vein thrombosis (DVT) (β =0.001787) but negative on dilated cardiomyopathy (DC) and cardiac valvular disease (CVD), as shown in Figure 1a and Table 1. These analyses indicate that ING/INB can exert pro-thrombotic effect potentially by agonizing AhR receptor. Furthermore, colocalization analysis confirms the QTL of AhR and the outcome (DVT) of ING/INB exposure share the same causal variant (H4=0.91) as shown in Figure 2 and Table S8, which corroborates the putative causal effect of the exposure (ING/INB→AhR) on DVT.

**Figure 2.**
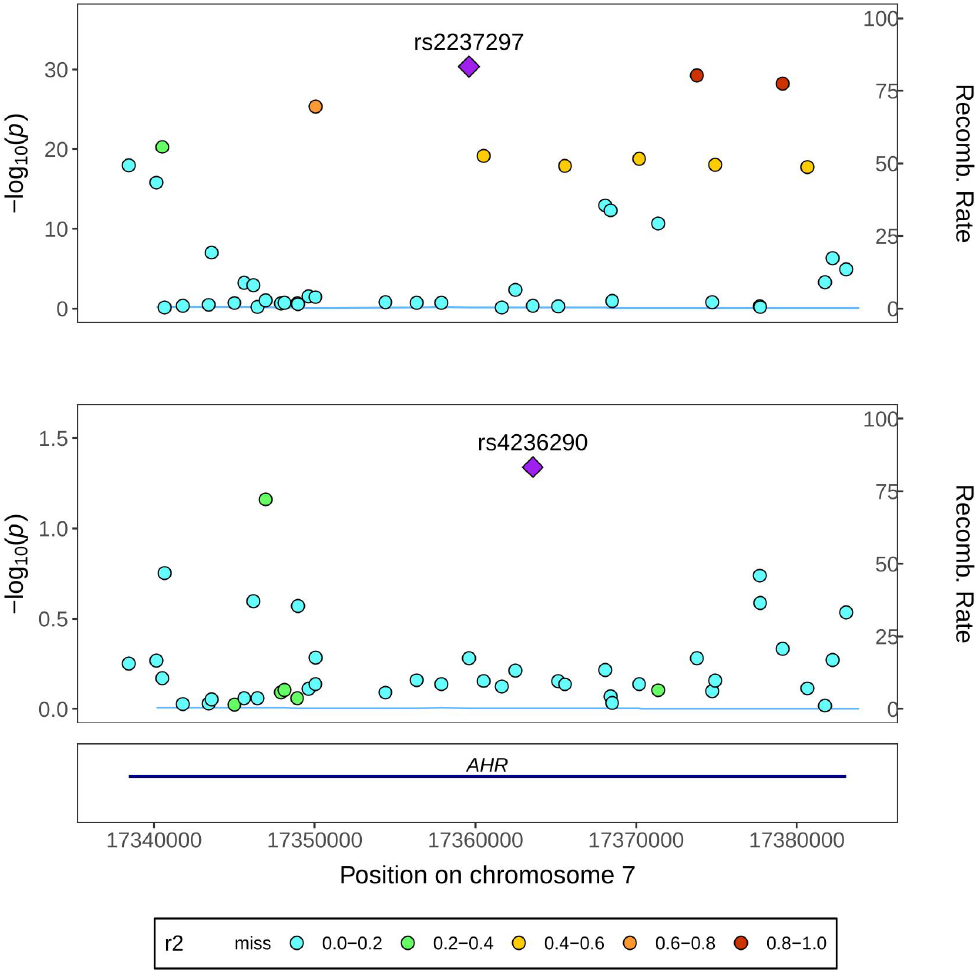
Colocalization analysis. Regional plots for the associations of AhR eQTL and DVT within ± 100 kb of AhR (GRCh37/hg19 by Ensembl, Chr7: 17,338,246-17,385,776).

In addition, MR displays a negative effect of FLT3 expression on arterial embolism and thrombosis (AET)(β =−0.7687) but positive on ischemic stroke (IS). These analyses suggest that INB can lead to AET potentially by antagonizing FLT3 receptor. Similarly, the expression levels of CDK4, 6 and 14 are inversely associated with pulmonary embolism (PE; β =−0.003284), congestive heart failure (CHF; β =−0.15), and hypertensive heart disease (HHD; β =−0.3768), respectively (Figure 1b and Table 1), which indicates ING/INB can cause PE, CHF or HHD by potentially antagonizing CDK receptors.

Sensitivity analyses were used to fix the issues of pleiotropy and heterogeneity, as shown in Table S9. Cochran’s Q-test and funnel plot showed no evidence of heterogeneity and asymmetry (Table S9). The MR-Egger intercept showed weak evidence of pleiotropy at the directional level (Table S9). Potential horizontal pleiotropy was not detected using MR-PRESSO global test (Table S9). In addition, the effect of each SNP on the overall causal estimates was verified by leave-one-out analysis (Table S9), indicating that all SNPs were calculated to make the causal relationship significant.

Of note, MR evidence for the causal effect of GSK-3β on 30 vascular disorders was not found at *P* level at 5×10^−5^. So, GSK-3 β was not included in this study.

### 3.2 Mediation effect

The mediation effect was studied by screening metabolites, immune cells and gut bacteria using two-sample MR. Urate was identified as a potential mediator between AhR and DVT (Table S7). As illustrated in Figure 3, the indirect effect of urate was estimated at 0.0001295 (95% CI: −0.0000008, 0.0002598), accounting for 7.25% of the total effect (0.001787).

**Figure 3.**
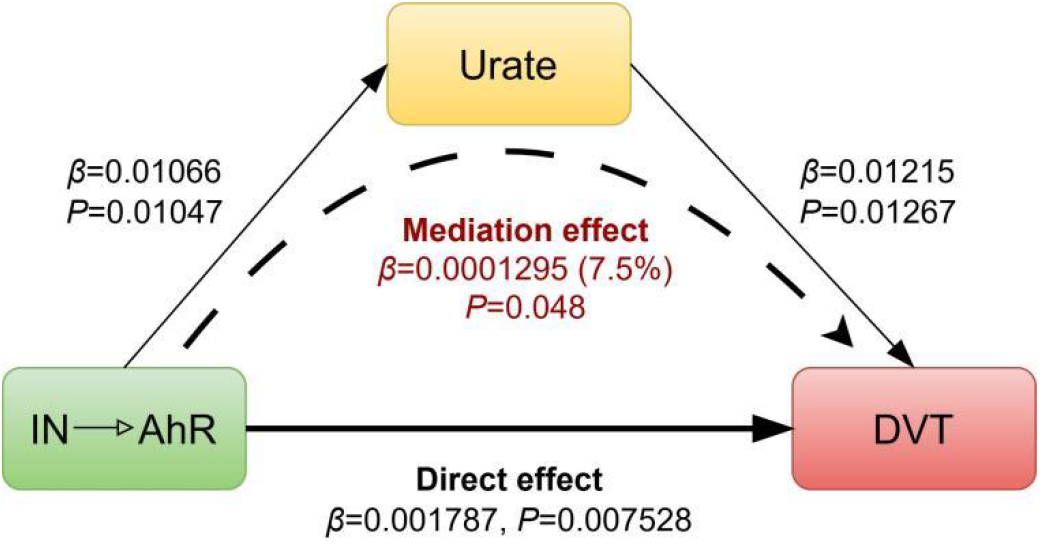
Result of mediation MR analysis (*P* = 5×10^−5^).

In contrast, the direct effect (0.001657) comprised 92.75% of the total effect, suggesting that the influence of AhR activation of ING/INB (exposure) on DVT (outcome) is primarily driven by non-mediated pathways. In addition, the casual association between urate and thrombosis is supported by clinical studies. For instance, serum uric acid at high levels is associated with an increased risk of VTE recurrence.^25^ Urate-lowering therapy mitigate the risks of hospitalized stroke and mortality in patients with gout.^26^

## 4. Discussion

This study investigated the causal relationships between 2 APIs of IN and 30 of vascular conditions. The causal effect of IN exposure on DVT outcome was identified, and urate mediates the effect with 7.5% of contribution. In addition, IN exposure is casually associated with multiple vascular risks, i.e., embolism and thrombosis including AET and PE, as well as heart diseases including CHF and HHD, which are driven by the antagonisic effect of INB on FLT3, CDK4, CDK6 and CDK14.

While IN has been recognized by TCM with an anti-inflammation effect on autoimmune inflammatory disorders, its side effect on vascular system has been typically observed in clinical trial and practice^4, 7, 8^. This work added MR evidence to these clinical observations and our understanding in this regard is proposed as follows.

### 4.1 Pharmacological duality of AhR pathway

The vascular toxicity of IN identified by this work can be explained by the biphasic pharmacological property of AhR. Through activating AhR with a EC_50_ value of 0.2 and 5 nM, respectively,^8, 12^ INB/ING can resume Th17/Treg balance, and consequently promoting IL-22 secretion and intestinal mucosal repair, which is the primary anti-inflammatory mechanism of IN,^3, 27^ as illustrated in Figure 4b. On the other hand, this anti-inflammatory effect can however shift to a certain pro-inflammatory response under specific pathological conditions, as a result, vascular toxicities would manifest. This transition involves AhR/CYP1A1 metabolic axis,^28^ which can be over-activated by INB/ING alongside ROS overproduction, leading to the assembly of NLRP3 inflammasomes (Figure 4c). Moreover, NLRP3 inflammasomes can be promoted by INB/ING in another ways. For example, by depleting available AhR, INB/ING can dampen the suppressive effect of AhR on NLRP3 inflammasome activity.^29^ Similarly, excessive activation of AhR by INB/ING would deplete endogenous AhR ligands via AhR/CYP1A1 axis, which would disrupt the homeostasis of indole derivatives and consequently induce oxidative toxicity.^30^ In fact, the presence of INB and ING in human urine at average concentrations of ~0.2 nm^12^ undercores the significance of physiological concentrations of INB and ING.

**Figure 4.**
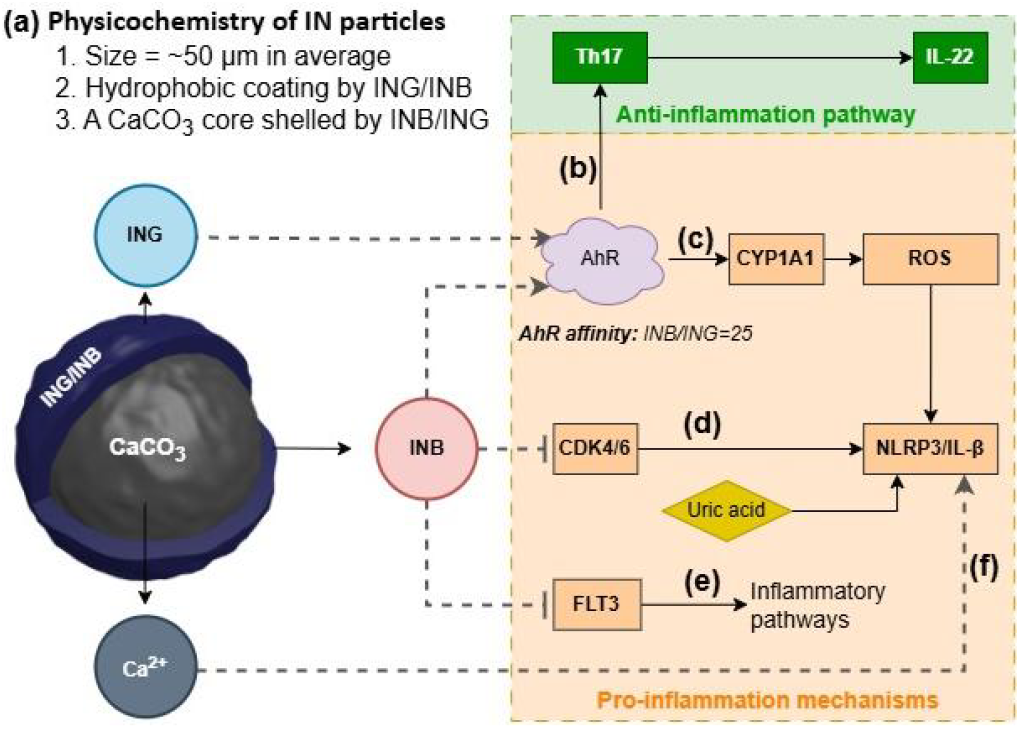
Physicochemistry and biology of IN. (a) Structural features, (b) anti-inflammatory and (c-f) pro-inflammatory mechanisms of IN.

In particular, INB would contribute more significantly to IN toxicity than its isomer ING. This is because that the 25-fold greater AhR affinity of INB than ING would amplify proinflammatory signaling,^8, 12^ although they share similar physicochemical properties (aqueous solubility ~40 μg/mL, LogP≈3.5).^31^

### 4.2 Pro-inflammatory pleiotropy of INB

INB is a potent inhibitor of FLT3 and CDKs.^17^ INB can cause inflammation through its inhibitory effect on FLT3 with an IC_50_ of 25 nM,^13^ and on CDKs with IC_50_ in the range of 50-100 nM,^13^ respectively (Figure 4d-e). Indeed, FLT3 inhibition can trigger an immediate global change in gene expression, leading to significantly upregulated innate immune and inflammatory pathways.^32–34^ In a similar line, CDK4/6 inhibition can activate NLRP3 inflammasomes and consequently facilitate IL-1β secretion.^35^ These reports indicate the pro-inflammatory pleiotropy of INB.

Interestingly, traditional folk practices of IN-based therapy involve washing IN with alcoholic drinks before oral administration for a purpose of reducing digestive system side effects. By analyzing an ethanolic extract, we found that INB was dominantly extracted yet not ING (data not shown). Both folk experience and laboratory analysis thus corroborate INB’s toxic potential. Notably, such difference in solubility between INB and ING in organic solvents suggests that they likely exhibit distinct profiles of bioavailability and toxicity, especially when considering the impact of food intake and the complex composition of the intestinal lumen. This certainly warrants a more detailed and thorough investigation.

### 4.3 Pro-inflammatory physicochemistry of particulate IN

Different from common TCM herbs, IN is processed using CaO and then presented as microscale particles with an inorganic core of CaCO_3_ and a hydrophobic surface due to the coating of INB/ING,^36^ as illustrated in Figure 4a. This unique structure can potentiate INB/ING-mediated inflammation through the following mechanisms. Ca^2+^ release can activate NLRP3 via the pathway of calcium sensing receptor,^37^ moreover, the resulted overload of Ca^2+^ can lead to oxidative stress,^38^ which then collectively facilitate the AhR/CYP1A1/ROS signaling. As a result, these events would finally converge to a hyperactivation of NLRP3 (Figure 4f).

In addition, IN has a hydrophobic surface because of INB/ING coating, which reduces the bioavailability and as a result, necessitates high dosing of IN, so, it would increase offtarget toxicity of IN. Besides, of particular concern is the typical use of CaO in traditional herbal processing,^39^ this highlights a demand to investigate safety risk of the interactions among calcium, herb chemicals and biological relevance.

### 4.4 Safety considerations for IN applications in TCM

IN has been used by TCM for UC treatment,^1, 27^ while UC are associated with higher levels of cardiovascular risks.^40, 41^ Considering the vascular risks of IN use, clinicians should exercise caution as IN may worsen clinical outcomes by increasing cardiovascular risks.^42^ This is especially critical for corticosteroid-treated UC patients, who already face significantly higher risks of venous thromboembolism and ischemic heart disease.^43^ Moreover, the chronic inflammatory nature of UC necessitates prolonged IN treatment, which raises a concern on the cumulative toxicity of ING/INB on vascular system. Indeed, it has been reported that IN treatment may have an undesirable association with an increase in pulmonary arterial systolic pressure in UC patients.^44^ In addition, IN is an anticancer agent for the treatment of acute myeloid leukemia (AML) in TCM,^17^ but, proinflammatory INB may potentially exacerbate AML because inflammatory signaling is typically involved in AML pathogenesis. Beyond the above, the synergy of metabolites such as uric acid in pro-inflammatory effect of IN should be taken into consideration (Figure 4d).

These risks of IN applications underscore a significant need to optimize traditional processing of IN to improve the safety profile while maintaining therapeutic efficacy. To address the challenges, three key improvements on the traditional processing of IN are proposed as follows: (1) reducing the percentage of INB by chemical engineering, (2) removing Ca^2+^ through innovations in herb formulating technology, and (3) developing drug delivery systems to improve therapeutic index.

### 4.5 Limitations of the study

This study has certain limitations. First, a range of other active components of IN such as alkaloids, nucleosides, amino acids, inorganic elements,^36^ were not included, although ING and INB occupy the majority of IN. For example, besides ING and INB, IN has many other indole alkaloids such as isatin, tryptanthrine, etc., which have been demonstrated with antiinflammation effect.^6^ Second, eQTLs may not capture posttranscriptional regulation and eQTL effects, which actually can vary by cell state/disease. Integration of pQTLs (protein QTLs) and use of eQTLs from disease-relevant tissues will improve our study. Lastly, MR findings suggest a potential causal relationship between genetic predisposition to an exposure and an outcome, yet they don’t demonstrate that therapeutic modifications of the exposure would significantly affect the outcome.

## 5. Conclusions

This study presents the first comprehensive investigation into the target-specific effect (AhR, CDK1-20, GSK-3β and FLT3) of herbal medicine IN on the vascular system. Utilizing MR analysis, we identified significant causal associations between IN exposure and multiple vascular conditions including DVT, PE and AET, as well as related heart diseases including CHF and HHD. Importantly, our findings identify urate as a key mediator in IN-induced DVT via AhR activation. These genetic insights highlight critical safety considerations for developing IN-based anti-inflammatory therapeutics and guide biomarker monitoring in related trials. In addition, this study proposed the potential role of pharmacological duality of AhR pathway, pleiotropy of INB, and physicochemistry of IN particles in the vascular toxicity of IN. Further study is in demand to focus on these aspects to advance the understanding of the vascular toxicity of IN.

## Author contributions

JL: Conceptualization, Data curation, Formal analysis, Funding acquisition, Investigation, Methodology, Project administration, Resources, Software, Validation, Visualization, Writing – original draft, Writing – review & editing. JCL: Methodology, Software, Writing – review & editing. HTX and ZPX: Writing – review & editing.

## Data availability

The data supporting this article are available in the manuscript or from the authors upon reasonable request.

## Acknowledgements

Jie Liu extends sincere gratitude to Fenji postdoctoral program in Shenzhen Bay Lab of Guang Dong Province of China. The authors acknowledge financial supports from the IDEATION Programme of the Hong Kong Science & Technology Parks Corporation (Partner-23-159). The methodological suggestions from Dr. ZHENG Ping are strongly appreciated.

